# Dangers of ACE inhibitor and ARB usage in COVID-19: evaluating the evidence

**DOI:** 10.1101/2020.03.25.20043927

**Authors:** Krishna Sriram, Paul A. Insel

## Abstract

**Background:** Concerns have been raised regarding the safety of Angiotensin Converting Enzyme Inhibitors (ACEIs) and Angiotensin Receptor Blockers (ARBs) in patients with COVID-19, based on the hypothesis that such medications may raise expression of ACE2, the receptor for SARS-CoV-2.

**Methods:** We conducted a literature review of studies (n=12) in experimental animals and human subjects (n=12) and evaluated the evidence regarding the impact of administration of ACEIs and ARBs on ACE2 expression. We prioritized studies that assessed ACE2 protein expression data, measured directly or inferred from ACE2 activity assays.

**Results:** The findings in animals are inconsistent with respect to an increase in ACE2 expression in response to treatment with ACEIs or ARBs. Control/sham animals show little to no effect in the plurality of studies. Those studies that report increases in ACE2 expression tend to involve acute injury models and/or higher doses of ACEIs or ARBS than are typically administered to patients. Data from human studies overwhelmingly imply that administration of ACEIs/ARBs does not increase ACE2 expression.

**Conclusion:** Available evidence, in particular, data from human studies, does not support the hypothesis that ACEI/ARB use increases ACE2 expression and the risk of complications from COVID-19. We conclude that patients being treated with ACEIs and ARBs should continue their use for approved indications.

## Introduction

There has been much recent debate regarding the use of ACE inhibitors (ACEIs) and Angiotensin Receptor Blockers (ARBs) in COVID-19 patients ^1-7^, thus prompting concern among patients and health care providers. The basis of this concern involves whether ACEIs/ARBs increase expression of ACE2, the primary cellular receptor for the SARS-CoV-2 virus, thereby possibly increasing severity of the infection. Published correspondences/letters such as those cited above typically make assertions on a relationship between ACEI/ARB use and ACE2 expression levels but lack a detailed, comprehensive breakdown of available data in humans and animals. The strength of the experimental data is thus unclear regarding ACEI/ARB use and ACE2 protein expression.

To address this issue, we have reviewed key literature by assessing studies conducted in experimental animals (primarily rats) and humans, in order to obtain a more comprehensive picture of what the available data convey. We focus our discussion on studies for which ACE2 protein data are available (either via direct measurement or inferred from ACE2 activity assays), as tissue ACE2 mRNA expression appears to only weakly correlate with protein expression, as shown from data in the Human Protein Atlas ^8^ (*proteinatlas.org*), results from human renal samples ^9^ and studies in experimental animals (discussed below [e.g., ^10,11^]).

We identified relevant studies by searches in Pubmed and Google Scholar for all available literature. Combinations of search terms were used to maximize the identification of relevant studies. Search terms included “ACE inhibitor”, “Angiotensin Receptor Blocker”, “ACE inhibition”, “ACE2”, “ACE2 expression”, “ACE2 protein expression”, “ACE2 activity”, “humans”, “patients”, “lung”, “heart”, “kidney”. We imposed no limits on when studies were performed. We manually curated each relevant hit to ensure the data were from articles with original research and/or meta-analysis and included quantitative/normalized ACE2 protein expression/activity and involved studies with tissues relevant to COVID-19/SARS-CoV-2 infection.

## Results

We focused our assessment on studies that evaluated ACE2 levels in tissues/cells involved in infection by SARS-CoV-1 or SARS-CoV-2. Data from 12 animal studies are summarized in **Table 1**, which shows the effect of ACEI/ARB use on ACE2 protein expression/activity in animals (primarily rats). We used conversion factors based on the principles of allometric scaling to evaluate doses in animals relative to their use in humans ^12^. For studies in which this was relevant, we used the HED (Human Equivalent Dose, assuming a 60 kg human ^12^) for doses of ACEI/ARBs. The conversion factor for rats was 6.2 (i.e., doses in mg/kg were divided by 6.2, then multiplied by 60 kg for a human equivalent dose) and 1.1 for pigs. We obtained recommended doses for humans using approved labelling from the Food and Drug Administration (FDA) website (*fda.gov*). These recommended human doses are provided in **Table 2**. For most of the drugs, the maximum doses indicated in **Table 2** are generally not administered to humans, such that treatment of animals with equivalent (or larger) doses, (especially in models of acute dosing, as in refs ^17^ and ^21^ discussed in **Table 1**) raise concern about their relevance to ACEI/ARB administration to patients.

**Table 1.** Studies in animals that have assessed ACE2 protein expression in response to ACEI/ARB treatment.

| Source | Study Details | Effect of ACEI / ARB on ACE2 |
| --- | --- | --- |
| Ferrario et al. <sup>10</sup> | Lewis rats were treated with losartan (an ARB) or lisinopril (an ACEI) 10 mg/kg/day (HED = 96.8 mg/day), for 20 days. ACE2 activity was measured in membranes from the renal cortex. | Lisinopril or Losartan treatment were both associated with increases in ACE2 activity but used in combination, did not produce this effect. |
| Ocaranza et al. <sup>13</sup> | Sprague Dawley (SD) rats were used in a myocardial infarction model, via coronary ligation. The ACEI Enalapril (10 mg/kg/day; HED = 96.8 mg/day) was administered for 8 weeks post- surgery. Plasma ACE2 activity was measured. | Enalapril increased plasma ACE2 by ~14% and ~36% in sham and MI animals, respectively. |
| Hamming et al. <sup>14</sup> | Renal ACE2 activity was assayed in Wistar rats, controls or on low sodium diet along with lisinopril (75 g/l) in drinking water for 3 weeks. | Renal ACE2 activity was unchanged with ACEI treatment in either group |
| Velkoska et al. <sup>15</sup> | ACE2 activity was assessed in kidneys from SD rats with subtotal nephrectomy and given ramipril (ACEI), 1 mg/kg/day; HED = 9.68 mg/day) for 10 days. | ACE2 activity in renal cortex and medulla was unchanged by ACEI treatment In control rats and increased ~50% with nephrectomy. |
| Han et al. <sup>16</sup> | . ACE2 protein expression in lungs was measured in SD rats with cigarette smoke-induced lung damage. Rats were treated with losartan (10 or 30mg/kg/day [HED = 96.8 or 290 mg/day]) for 6 months. | ACE2 expression was unchanged in control rats by either dose of losartan. Animals exposed to cigarette smoke had reduced ACE2, which losartan treatment restored. |
| Wösten-van Asperen et al. <sup>17</sup> | SD rats were used in a LPS-induced model of Acute Respiratory Distress Syndrome (ARDS). Rats were given losartan (2.5 mg/kg/h during 4h of ventilation [HED = 96.8 mg]),. ACE2 protein expression in the lung was measured 24h after inducing ARDS with LPS. | Losartan administration decreased ACE2 activity in control animals (unclear if/how statistics were performed). After induction of ARDS, ACE2 levels decreased and were restored to normal by losartan. |
| Burrell et al. <sup>18</sup> | ACE2 activity and protein expression were assayed in tissues from SD rats 28 days after subtotal nephrectomy and which received ramipril (an ACEI, 1 mg/kg/day [HED = 9.68 mg/day). | Ramipril had no effect on ACE2 in cardiac or renal (cortex or medullary) tissue. ACE2 activity was reduced by nephrectomy; ramipril restored ACE2 activity to control levels in renal cortex, but not in medullary or cardiac tissue. |
| Burchill et al. <sup>19</sup> | ACE2 protein expression was assessed in cardiac tissue in a Myocardial Infarction (MI) model in SD rats. Ramipril (1 mg/kg/day, HED = 9.68 mg/day) and Valsartan (ARB, 10 mg/kg/day; HED = 96.8 mg/day) were given for 28 days post-coronary artery ligation. | ACE2 expression was not altered but may have decreased in viable myocardium border or infarct zones, (unclear statistical analysis). |
| Yang et al. <sup>11</sup> | Spontaneously Hypertensive rats were treated with enalapril (15 mg/kg/day; HED = 145.2 mg/day) for 4 weeks. Cardiac ACE2 protein expression was measured. | ACE2 mRNA expression was increased but ACE2 protein expression did not change with ACEI treatment |
| Zhang et al. <sup>20</sup> | . Cardiac ACE2 protein was assayed from SD rats with cardiac remodeling from aortic constriction and treated with losartan (30 mg/kg/day; HED = 290.3 mg/day) or enalapril (20 mg/kg/day; HED = 193.5 mg/day) for 20 days, starting 4 days after surgery. | ACE2 cardiac protein expression was increased (~3-fold) with both drugs in rats with cardiac remodeling; data were not provided for animals with sham surgery. |
| Li et al. <sup>21</sup> | SD rats underwent LPS-stimulated lung injury. Simultaneous with intravenous injection of LPS, animals were administered captopril (50 mg/kg [HED = 483.9 mg]). After 8h, samples were collected and ACE2 protein expression was measured in the lung. | ACE2 protein expression was elevated in the lung, in control rats (~35%) and those with LPS-induced lung injury (~ 2-fold). |
| Wang et al. <sup>22</sup> | Pigs were used to study effects of ACEIs on cardiac arrest and resuscitation. Enalapril (0.2 mg/kg; HED = 10.9 mg) was perfused for 30 min, followed by surgery. Myocardial ACE2 protein expression was assayed in samples collected 6h post- surgery. | Compared to saline-infused controls, enalapril did not increase ACE2 levels; enalapril was not administered to sham rats. |

**Table 2.** FDA-recommended doses of the ACEIs/ARBs discussed in Table 1, along with pharmacokinetics data in humans and rats.

| **Drug** | **Typical initial daily adult dose** | **Maximum daily adult dose** | **C<sub>max</sub> in humans** | **C<sub>max</sub> in rats** |
| --- | --- | --- | --- | --- |
| Losartan | 50 mg | 100 mg | 250 ng/ml, for 50 mg dose <sup>23</sup> | 1260 ng/ml, for 10 mg/kg dose; HED = 96.8 mg <sup>24</sup> |
| Enalapril | 5 mg | 40 mg | 62 ng/ml for 10 mg dose <sup>25</sup> | 1329 ng/ml for 15 mg/kg dose; HED = 145.2 mg <sup>26</sup> |
| Lisinopril | 10 mg | 40 mg | 38 ng/ml for 10 mg <sup>27</sup> | 6400 ng/ml for 5 mg/kg dose; HED = 48.4 mg <sup>28</sup> |
| Ramipril | 2.5 mg | 20 mg | 52.2 ng/ml at 10 mg dose <sup>27</sup> | 50.45 ng/ml for 1 mg/kg dose; HED = 9.68 mg <sup>28</sup> |
| Captopril | 50 mg | 450 mg | 1580 ng/ml for 100 mg dose <sup>30</sup> | 9200 ng/ml for 30 mg/kg dose; HED = 290.4 mg <sup>30</sup> |
| Valsartan | 80 mg | 320 mg | 2000 ng/ml for 80 mg dose <sup>23</sup> | ~3700 ng/ml for 4.71 mg/kg dose; HED = 45.6 mg <sup>31</sup> |

In summary, 3 studies ^10,13,21^ in animals reported an increase in all treatment groups, of ACE2 protein expression/activity with ACEI/ARB treatment but in one such study,^10^ combined ARB/ACEI treatment did not show this effect. The other two studies reported relatively small effects (< 40% increase in ACE2 protein expression in control/sham animals). All of these studies used high doses of ACEIs/ARBs, as noted above. By contrast, 6 studies ^11, 14, 16, 18, 19, 22^ found little or no change in ACE2 expression with ACEI/ARB treatment. In 3 studies ^15, 17, 21^, treatment with ACEI/ARB had no effect or a decrease in ACE2 protein expression/activity in control/sham animals; increased ACE2 expression was only observed following experimental exposure, such as lung injury, myocardial infarction, etc. In nearly all studies that reported an increase in ACE2, doses of ACEIs/ARBs used were greater than equivalent doses typically administered to patients. We identified only one study ^15^ in which ACEI/ARB use increased ACE2 protein expression at doses typically used to treat patients, albeit these changes only occurred after exposure to acute injury (subtotal nephrectomy). Overall, the studies with experimental animals do not provide consistent evidence for an effect of ARB/ACEI administration on ACE2 protein expression, especially in contexts that model drug administration in humans.

A challenge with assessing the animal studies in **Table 1** is that while doses were provided for the drugs, pharmacokinetic studies were not performed to ascertain peak serum concentrations (C_max_) and Area Under the Curve (AUC), which would allow more precision in evaluating the translational relevance of findings. However, data from other studies provide insight into the pharmacokinetics in rats, at doses comparable to those used in the studies in **Table 1. Table 2** shows estimates for C_max_ for the drugs used in studies in **Table 1**, in humans (at therapeutic doses, as indicated) and rats (at doses comparable to those in **Table 1**). The differences in AUC for these drugs between humans and rats mirror differences in C_max_; we refer readers to the indicated references for further details. With the exception of a study in a nephrectomy model^15^, animal data (including all data for control animals) that show a relationship between ACE2 expression and use of ACEIs/ARBs involved drug treatments associated with plasma concentrations far in excess (often more than one order of magnitude) of values normally seen in patients. Absent exposure-response studies with more clinically appropriate drug concentrations, we conclude that the bulk of the animal data regarding ACE2 expression with ACEI/ARB administration has dubious translational relevance.

Evidence of changes in ACE2 protein in human subjects/patients (**Table 3**) are derived from studies that assessed ACE2 protein concentration or enzymatic activity in urine or serum/plasma. Of the 11 studies summarized in **Table 3**, 7 showed no effect of ARB/ACEI use on ACE2 protein levels in any conditions/patient grouping. One study ^37^ documented a small increase in serum ACE2 attributable to use of ACEIs among Type-1 diabetic patients but found no effect from use of ARBs. Another study ^34^ found a slightly larger proportional decrease in urinary ACE2 in patients with Type-2 diabetes using ACEIs/ARBs but did not distinguish between effects from use of ACEIs or ARBs. In one study ^33^ the investigators observed that subjects using the ARB olmesartan had increased ACE2 levels, but several other ARBs and ACEIs had no effect. In one study ^39^, ACEI use had no effect in control, stage 3-5 chronic kidney disease patients or those on dialysis; ARB only had a small effect in patients on dialysis. Besides these quantitative data, another study used immunohistochemical analysis to assess ACE2 protein expression in the kidneys and found no ACEI-dependent effect^43^. Together, these 12 studies in humans imply a lack of association between ACE2 protein expression and the use of ARBs or ACEIs and support the idea that ACEIs/ARBs are unlikely to raise ACE2 or be harmful in the context of COVID-19 infection.

**Table 3:**
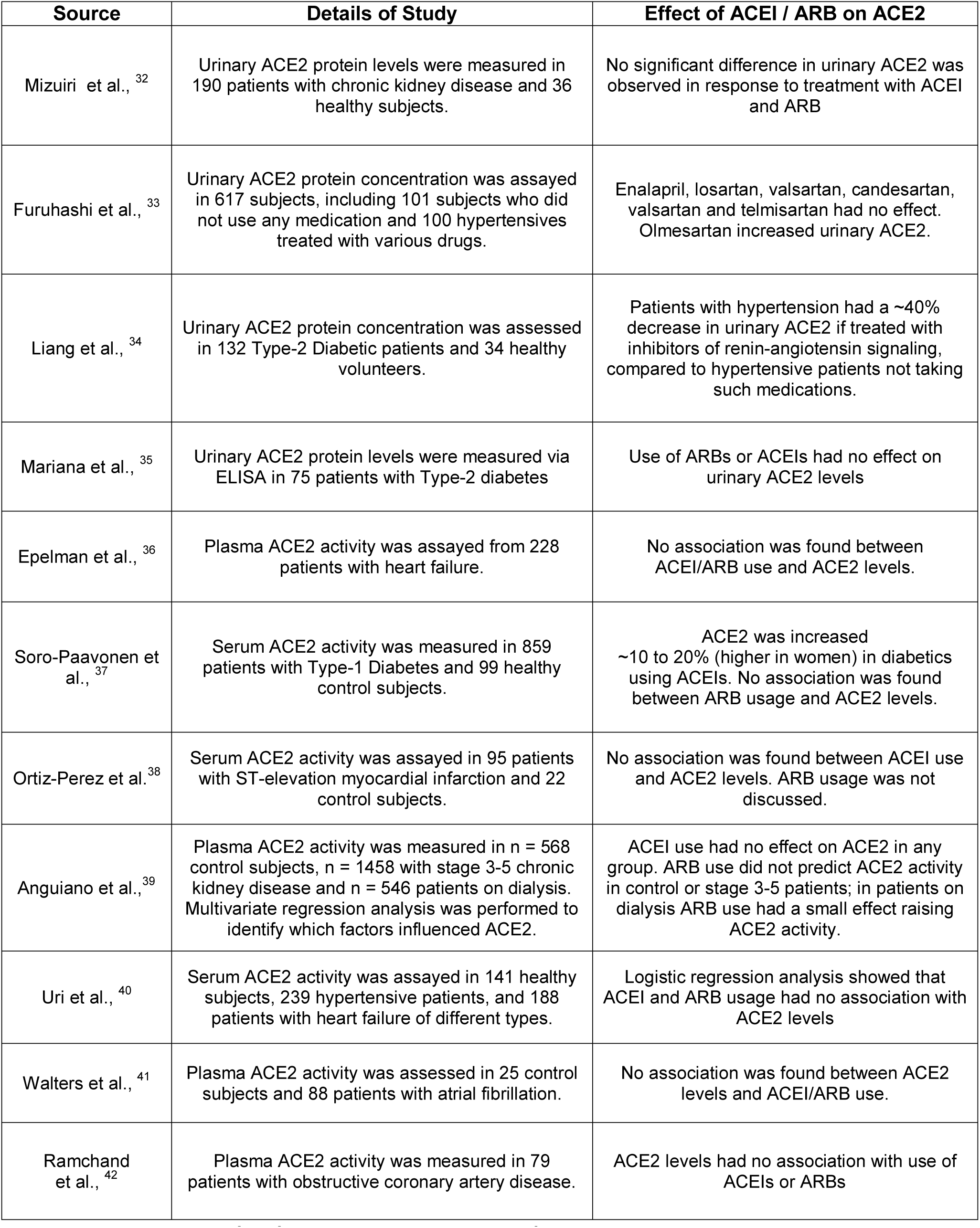
Studies in humans of the relationship between ACEI/ARB use and ACE2 protein expression. Entries are ordered chronologically, first for studies in urine and then for studies in circulating ACE2.

## Discussion

What would constitute strong, supportive evidence for the hypothesis that ACEI/ARB usage is a risk factor in the setting of SARS-CoV2 infection? Findings to help support that hypothesis would include: 1) Replication of a prominent effect in multiple animal studies and models; 2) Evidence that tissues with low expression of ACE2 have prominent increases in its expression and activity following ACEI/ARB treatment; 3) Data documenting that increases in ACE2 expression in response to ACEI/ARB treatment enhance the ability of the SARS-CoV-2 virus to infect cells; 4) Findings from human studies of statistically significant relationships between ACEI/ARB usage and ACE2 expression/activity; 5) Epidemiological data showing that COVID-19 patients administered ACEIs/ARBs have increased morbidity and mortality, ideally with a dose-response relationship for such outcomes.

How well do the available data provide such evidence? 1) The hypothesis that ACE2 expression increases with ACEI/ARB use is not supported by the plurality of available data from animal studies. 2) There is no available evidence for de novo expression of ACE2 expression in response to ACEIs/ARBs in tissues with low expression. 3) The affinity of SARS-CoV-2 for ACE2 is very high, ∼4-fold greater than SARS-CoV-1 ^44^ or higher; some studies suggest an order-of-magnitude higher affinity for SARS-CoV-2 ^45^. It is unclear if small or modest perturbations in ACE2 expression impact the infectivity of SARS-COV-2. Moreover, ACE2 levels may decrease with age ^46,47^ and diabetes ^48^ yet elderly/diabetic subjects are more vulnerable than younger individuals to COVID-19 ^49^. Modest changes in ACE2 expression (<2-fold, from most preclinical data discussed above) may not meaningfully impact on the high infectivity of SARS-CoV-2 in host tissues. 4) Data from human studies suggest that treatment with ACEIs/ARBs produces little or no effect on urinary or circulating ACE2 levels. As a caveat, changes in ACE2 levels in serum or urine may not reflect changes in tissue. 5) Epidemiological studies are largely unavailable in matched groups of patients infected with SARS-CoV-2, who have or have not been taking ACEIs/ARBs. One preliminary study, ^50^ examined 28 patients with severe COVID-19 and 18 patients with mild disease, all of whom also had hypertension. ARB use was associated with a reduction in risk of severe COVID-19 disease, morbidity and mortality, a result that contradicts the hypothesis that ARB/ACEI use is harmful. However, only small numbers of patients have been assessed.

Based on the data summarized above, we conclude that current evidence, especially from human studies (**Table 3**), does not support the idea that treatment with ACEIs or ARBS produces pathophysiologically relevant increases in ACE2 protein abundance. The hypothesis that the use of these drugs increases SARS-CoV-2 virus infectivity and/or severity of COVID-19 is therefore not supported by the available evidence. It would thus seem prudent for patients to continue receiving these medications, as recently recommended by multiple health associations^2^ and other publications ^51^.

## Study Highlights

### What is the current knowledge on the topic?

Debate exists regarding the evidence for ACE inhibitor/ARB usage and expression levels of ACE2 protein. This is of concern in COVID-19 because ACE2 is the receptor for the SARS-CoV-2 virus.

### What question did this study address?

We critically evaluated the evidence in the literature regarding the topic indicated above.

### What does this study add to our knowledge?

This is the most comprehensive assessment of the evidence behind the hypothesis that ACEIs/ARBs may be harmful in COVID-19, due to their up-regulation of ACE2 expression. We find that there is little evidence to justify these concerns. In particular, data from studies with human subjects refute the presence of a meaningful effect of ACE inhibitors or ARBs on the expression of ACE2 protein.

### How might this change clinical pharmacology or translational science?

Concern has been raised regarding the administration of ACE inhibitors or ARBs to patients during the COVID-19 pandemic. Several articles have suggested that such drugs may be harmful, causing understandable confusion among patients and providers. We conclude that these concerns are not supported by available data and that clinicians should continue to prescribe and patients should continue to use these drugs for approved indications.

## Data Availability

There are no underlying data to report, this is a review of data in the literature, cited where appropriate.

## Author Contributions

KS compiled and analyzed data and wrote the manuscript. PAI wrote and edited the manuscript.

